# Transmission routes of SARS-CoV-2 among healthcare workers of a French university hospital in Paris, France: a case-control study

**DOI:** 10.1101/2020.10.30.20223081

**Authors:** Adrien Contejean, Jérémie Leporrier, Etienne Canouï, Jacques Fourgeaud, Alice-Andrée Mariaggi, Fanny Alby-Laurent, Emmanuel Lafont, Lauren Beaudeau, Claire Rouzaud, Fabienne Lecieux, Agnès Greffet, Anne-Sophie L’Honneur, Jean-Marc Tréluyer, Fanny Lanternier, Anne Casetta, Pierre Frange, Marianne Leruez-Ville, Flore Rozenberg, Olivier Lortholary, Solen Kernéis

## Abstract

In this case-control study on 564 healthcare workers of a university hospital in Paris (France), contacts without protection with COVID-19 patients or with colleagues were associated with infection with SARS-CoV-2, while working in a COVID-dedicated unit, using public transportation and having children kept in childcare facilities were not.

## Introduction

Effective protection of healthcare workers (HCW) against severe acute respiratory syndrome coronavirus-2 (SARS-CoV-2) requires precise assessment of the respective role of in-hospital and out-of-hospital exposures on transmission routes in this at high-risk population.

We previously published an observational multicenter cohort study on HCW infected by SARS-CoV-2 during the first French coronavirus disease-19 (COVID-19) breakthrough in 2020 spring[1]. Only 20% of the HCW screened positive for SARS-CoV-2 reported close contact with a suspected or confirmed COVID19 patient without Personal Protective Equipment (PPE), and 78% were not regularly posted in a COVID-19-dedicated unit. On the other hand, 54% declared frequent close contacts during occupational activities with colleagues without protection. We were however unable to compare positive HCW to a robust control group gathering personnel not infected with SARS-CoV-2. Diagnosis sensitivity of reverse transcriptase polymerase chain reaction (rtPCR) on nasopharyngeal swab for COVID-19 is imperfect[2] and serological assessment was not available at this time. IgG serological test has since been proven to be reliably associated with a COVID-19 past-infection[3].

We present here a case-control study which aimed to compare COVID-19 positive and negative HCW regarding their occupational activity, symptoms and in-hospital and out-of-hospital exposures to SARS-CoV-2, to determine the respective roles of different SARS-CoV-2 transmission routes.

## Material and methods

### Patients and design

This case-control study was led among HCW of a 2,100-bed tertiary-care university hospital (AP-HP. Centre, Université de Paris) located in central Paris, France, employing 13,278 personnel. From 24^th^ February, to 10^th^ April 2020, symptomatic staff were referred to dedicated on-site screening centers where trained medical staff collected a nasopharyngeal swab for SARS-CoV-2 rtPCR. HCW who tested positive on rtPCR were included as cases (HCW+). For each confirmed case, we included a control symptomatic HCW tested for SARS-CoV-2 on the same day, who had a negative rtPCR and a negative serological assessment performed at least 1 month after symptoms onset (HCW-). Both cases and controls were questioned in detail on their professional activity, symptoms, occupational and non-occupational exposures to SARS-CoV-2 immediately after screening[1]. During this period, social distancing measures were implemented in France. On March 12^th^ all school and childcare facilities closed, except for children of hospital staff, and a nationwide lockdown started on March 17^th^. Lift of containment measures occurred on May 11^th^.

### Virology methods

SARS-CoV-2 rtPCR technique has been described elsewhere[1]. SARS-CoV-2 serology was determined by the Abbott® SARS-CoV-2 IgG assay, a chemiluminescent microparticle immunoassay intended for the qualitative detection of IgG antibodies to SARS-CoV-2.

### Statistical analysis

Continuous variables are presented as median (interquartile range) and categorical variables as number (percentage). Fisher exact tests were used for comparisons of qualitative variables and Mann-Whitney tests for quantitative variables. All tests were 2-sided with a .05 value for significance. Factors associated with SARS-CoV-2 infection were assessed using multivariate logistic regression models. To account for the impact of the lockdown on community exposures, we considered two periods before and after March 22^nd^, 2020 (March 17^th^ [date of national lockdown] + mean incubation period of 5 days). For each period, we first entered all exposures with a p-value < 0.40 in a multivariate model then used a backward stepwise selection procedure (removal criteria: p□>□0.05) to build the final model. Statistical analyses were performed using R-software (3.3.2, R Foundation for Statistical Computing, Vienna, Austria).

## Results

Between February 24^th^ and April 10^th^, 2020, 1344 symptomatic HCW were screened for SARS-CoV-2 by rtPCR on a nasopharyngeal swab. Among them, 373 had positive rtPCR results (28%), 336 (90%) completed the questionnaire, and were included as cases (HCW+). Among 338 matched HCW with negative rtPCR, 247 (73%) had a serological assessment, and 228 (92%) tested negative. This group of 228 HCW with both negative rtPCR and serology constituted the control group (HCW-).

Exposures reported by cases and controls are displayed in the Table. Cases and controls where comparable in terms of age, sex and professional category. Cases presented more frequently with anosmia or ageusia and with asthenia, fever, muscle pain, dyspnea, headaches and diarrhea than controls. Frequency of cough or rhinorrhea did not differ between groups.

In the univariate analysis (table), occupational activities with direct patient facing or assignment to a COVID-19-dedicated unit were similar in cases and controls. However, cases reported more close contacts with suspected or confirmed COVID-19 patients without PPE. Controls declared better compliance to mask wearing during occupational activities or in the presence of colleagues. Out-of-hospital exposures did not differ between cases and controls except for mask wearing outside home. In particular, using public transportation, having a child at home or a child kept out of the household were not associated with a significant increased risk of COVID-19.

Multivariate analysis showed that in the pre-lockdown period, close contacts with colleagues without protection (OR 2.58 [1.49-4.60]) was independently associated with an increased risk of COVID-19 in HCW, whereas systematic mask wearing outside of home was found independently protective against COVID-19 in this population (OR 0.43 [0.21-0.85]). Conversely, in the post-lockdown period, only close contacts with suspected or confirmed COVID-19 patients without PPE (OR 3.87 [1.73-9.89]) was independently associated with an increased risk of COVID-19 in HCW.

## Discussion

This case-control study among HCW highly exposed to SARS-CoV-2 suggests that PPE and especially medical mask is efficient in preventing COVID-19 transmission in HCW, including its systematic use outside hospital and home. To our knowledge, it is the first study to report both occupational and non-occupational exposures associated with SARS-CoV-2 infection in HCW since previous reports mainly focused on professional exposures and did not use IgG serology to contribute ruling-out COVID-19 in controls[4,5].

Unsurprisingly, symptoms were significantly different between cases and controls. Anosmia and ageusia appeared to be strongly associated to SARS-CoV-2 infection as reported elsewhere[6,7], whereas cough or rhinorrhea were not. Clinical scores including number of symptoms statistically associated with COVID-19 have been developed and might be a useful tool to quickly sort symptomatic HCW before results of SARS-CoV-2 testing[7].

Occupational risk factors for COVID-19 in HCW are dominated by exposure to suspected or confirmed COVID-19 patients without PPE or repeated contacts with numerous colleagues without medical mask. Indeed, SARS-CoV-2 infectiousness starts up to two days before symptoms onset[8], thus strict compliance to universal masking and social distancing measures are critical to prevent SARS-CoV-2 transmissions from asymptomatic patients or colleagues. Moreover, adherence to PPE during patient care is crucial to prevent COVID-19 infection in HCW, as we previously suggested[1]. Conversely, in our cohort, direct patient care was not associated with an increased risk of COVID-19 (including in COVID-19-dedicated wards), as long as HCW wear PPE. Of note, PPE supplies were immediately and fully available in our center, which was not the case in all French healthcare settings. Compliance to protective measures may also have been better among highly trained HCW of COVID-19-dedicated units. Recently, a large cohort study on 99,795 HCW suggested that frontline HCW were at increased risk of COVID-19 compared to community individuals, especially in case of exposition to patients with inadequate PPE[4].

Analysis of non-occupational exposures suggest that wearing a mask outside home may provide protection against COVID-19. The specific effect of mask wearing in the community is controversial. Indeed, HCW who reported wearing a mask outside home in our cohort (17%) were also probably more cautious regarding social activities and other suspected sources of SARS-CoV-2, which were not assessed in the questionnaire. Community use of facemasks have failed to demonstrate efficiency in prevention of influenzae transmission in a meta-analysis[9]. In Hong-Kong, a study suggested that the number of COVID-19 clusters were reduced when universal masking was recommended[10], and a study conducted in the USA concluded that mask wearing mandatory permitted to reduce daily COVID-19 growth rates[11]. However, to our knowledge, no study with high level of evidence has been published yet on that question.

As suggested in our first report[1] and here confirmed by the case-control study, HCW who reported to have children kept outside the family home did not have a higher risk of COVID-19 infection. However, childcare facilities that remained open for HCW’s children during the national lockdown gathered a limited number of children simultaneously (<10). This question of SARS-CoV-2 transmission from children is highly debated since the beginning of the pandemic, but accumulating data suggest that children are not significant drivers for COVID-19 pandemic[12]. Therefore, our results suggest that keeping schools and childcare facilities open for HCW is acceptable especially in case of second containment period, when HCW should be fully available at hospital.

We acknowledge several limitations, in particular recall bias, but cases and controls were interrogated prospectively and shortly after PCR assay. Additionally, our questionnaire included declarative data maybe not fully compressive regarding SARS-CoV-2 exposures.

In conclusion, profession category, direct patient care or assignment to a COVID-19-dedicated unit were not associated with COVID-19 infection in our cohort of HCW. Conversely, close contacts with suspected or confirmed COVID-19 patients without PPE or multiplication of contacts with colleagues without mask may be risk factors for COVID-19 infection in HCW. Systematic mask wearing outside home was associated with a reduction of this risk during the pre-lockdown period.

## Data Availability

All data referred in the manuscript are available.

## Authors’ Contributions

AC, JL, OL and SK designed the study and drafted the paper. AC, JL, ML, FR, OL and SK contributed to data analysis and interpretation. All authors critically revised the manuscript for important intellectual content and gave final approval for the version to be published. SK had full access to all the data in the study and had final responsibility for the decision to submit for publication.

## Ethical Considerations

This study was approved by the Ethical Review Committee for publications of the Cochin University Hospital (number AAA-2020–08012). According to French policy, a non-opposition statement was obtained for all participants, meaning that all had received written detailed information on the objectives of the study and were free to request withdrawal of their data at any time.

## Acknowledgments

The authors warmly thank medical students involved in data collection: Laurence Clastres, Mathilde Lehmann, Aline Pellegrini, Marine Sisouvan, Ilana Slotine, Diem Soubou and Abigaëlle Vergnet, the following physicians and nurses who actively contributed to the screening: Claire Aguilar, Caroline Charlier, Gérard Chéron, Claudine Duvivier, Muriel Fortier, Remy Gauzit, Béatrice Grandordy, Marc Lecuit, Léonie Meyer, Gabrielle Paluszek, Perrine Parize, Guillemette Thin and Christine Vinter.

---

**Table:**
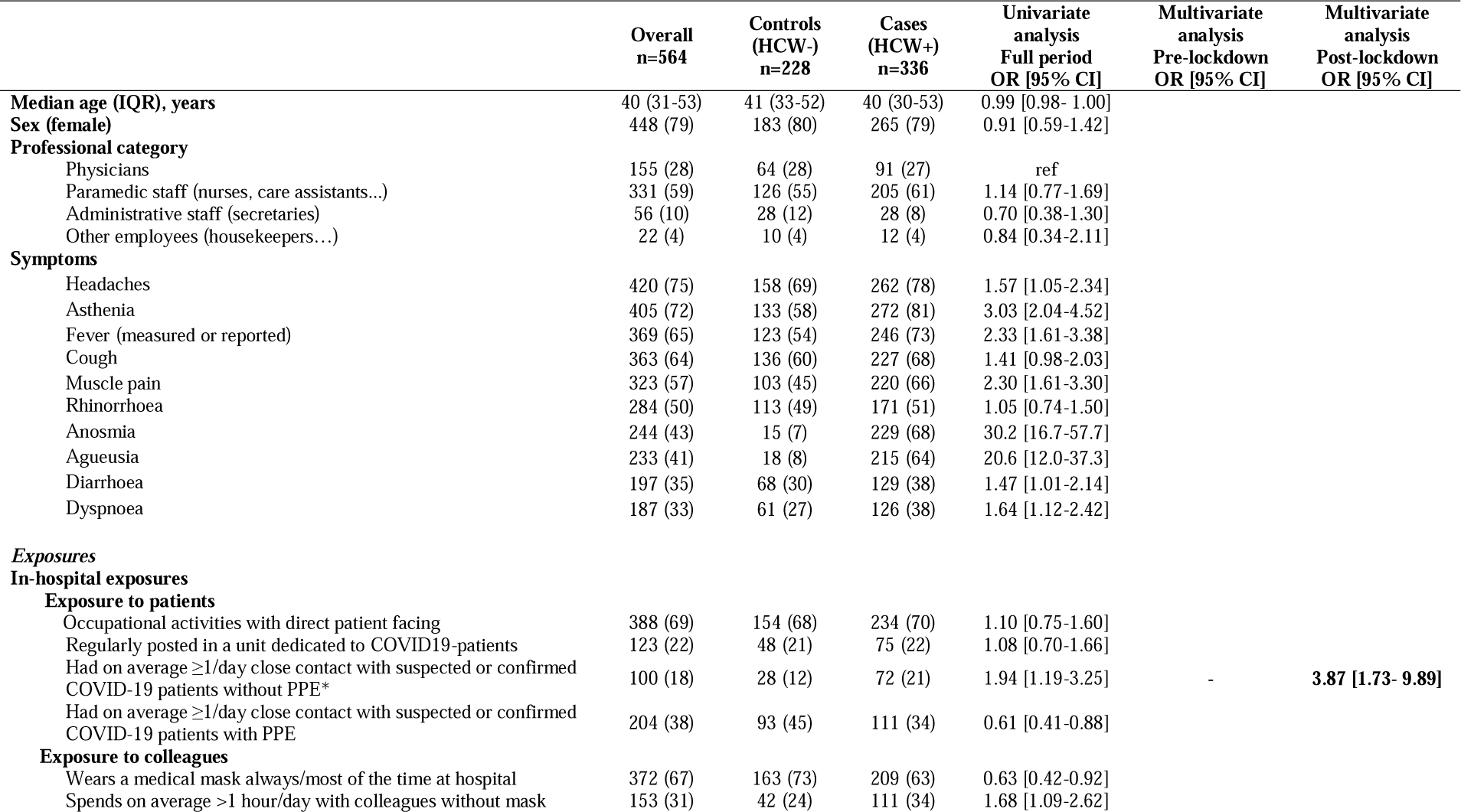
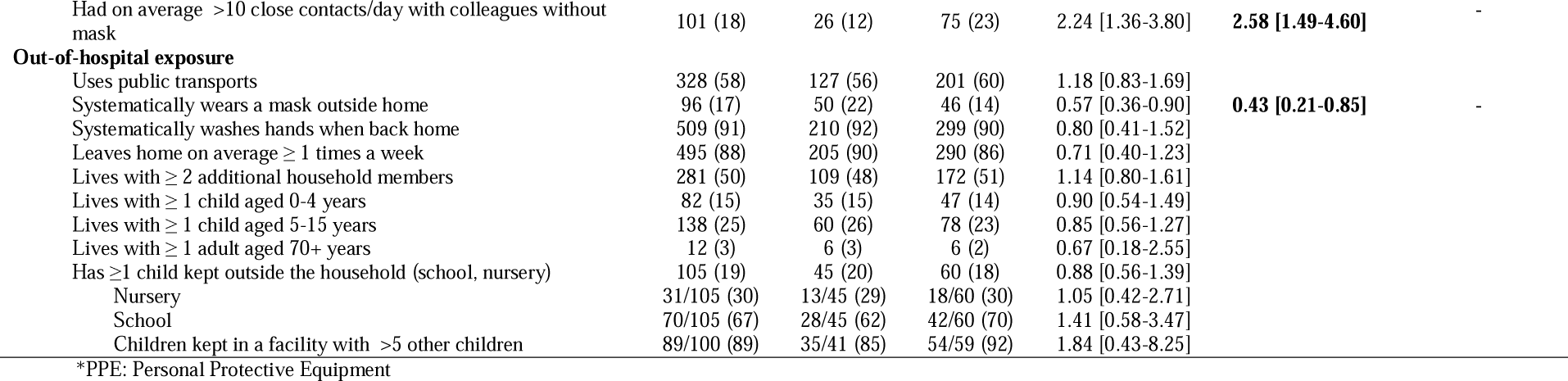
Cases and controls comparisons regarding demography, profession, symptoms and occupational or out-of-hospital exposures.

